# Risk factors related to hepatic injury in patients with corona virus disease 2019

**DOI:** 10.1101/2020.02.28.20028514

**Authors:** Lu Li, Shuang Li, Manman Xu, Pengfei Yu, Sujun Zheng, Zhongping Duan, Jing Liu, Yu Chen, Junfeng Li

**Author notes:** Corresponding author: Yu Chen, Difficult & complicated liver diseases and artificial liver center, Beijing You’an Hospital, Capital Medical University, Beijing, 100069, China., Junfeng Li, Institute of Infectious Diseases, Department of Infectious Diseases, The First Hospital of Lanzhou University, 1 Donggangxi Road, 730000, Lanzhou, China. Co-first authors:Lu Li and Shuang Li contributed equally to this work.

## Abstract

**Aims:** Corona virus disease 2019 (COVID-19) has rapidly become the most severe public health issue all over the world. Despite respiratory symptoms, hepatic injury has also been observed in clinical settings. This study aimed to investigate the risk factors involved with hepatic injury in the patients with COVID-19.

**Methods:** A total of 85 hospitalized patients who were diagnosed with COVID-19 in Beijing You’an Hospital were retrospectively analyzed. According to liver function, they were divided into ALT normal group (n=52) and ALT elevation group (n=33). Clinical features and laboratory data were compared between the two groups. The independent risk factors for liver injury were analyzed.

**Results:** There were 33 patients with hepatic injury in our study, accounting for 38.8% (33/85). The patients in ALT elevation group were older than those in ALT normal group. The levels of lactic acid, CRP, myoglobin, and neutrophils were significantly higher in ALT elevation group. The lymphocytes and albumin were significantly lower in ALT elevation group. The proportion of severe and critical patients in ALT elevation group was significantly higher. Multivariate logistic regression analysis showed CRP ≥20 mg/L and lymphocyte count< 1.1×10^9/L were independently related to hepatic injury.

**Conclusions:** Lymphopenia and CRP may serve as the risk factors related to hepatic injury in patients with COVID-19, which might be related to inflammatory cytokine storm in liver injury. Early detection and timely treatment of hepatic injury in patients with COVID-19 are necessary.

## Introduction

The corona virus disease 2019 (COVID-19), a severe public health issue in the world, is a newly emerging infectious disease caused by a novel coronavirus SARS-CoV-2, which may develop to acute respiratory distress syndrome or multiple organs failure^1-3^. It’s reported that this novel coronavirus is identified to the species of Severe Acute Respiratory Syndrome related-Coronavirus (SARSr-CoV) which belongs to the beta-coronavirus family and share 79.6% sequence identity to SARS-CoV ^4^. Based on the phylogeny, taxonomy and established practice, it was officially named as SARS-CoV-2^5^, and the disease caused by SARS-CoV-2 was named as COVID-19 ^6^.

Despite atypical pneumonia as the primary symptom, liver dysfunction has also been observed in many clinical cases^7-9^, indicating a possibility for the patients with COVID-19 may cause hepatic injury. Except that part of patients receiving lopinavir/ritonavir during hospitalization, which can cause drug-induced hepatic injury^10^, the underlying mechanisms for hepatic injury in patients with COVID-19 are still unclear. And there are few of clinical research regarding on this issue and it remains controversial now.

Therefore, the present study aimed to investigate the changes of liver injury in the patients with COVID-19 retrospectively, and analyzed the clinical features and laboratory data to seek the possible risk factors of liver injury in patients with COVID-19. The findings provide new clues and data for the mechanisms of liver injury in patients with COVID-19.

## Methods

### Study design and patient selection

From January 21st, 2020 to February 29th, 2020, a total of 85 consecutive patients with COVID-19 were admitted and treated in Beijing You’an Hospital, Capital Medical University, Beijing, China. The diagnosis criteria of COVID-19 are based on the detection of nucleic acid, the epidemiological history, clinical feature or laboratory testing and imaging^11^.

The clinical classifications are as follows: (1) Mild: only mild symptoms, no sign of pneumonia in imaging. (2) Ordinary: with fever, respiratory tract symptoms, and imaging shows pneumonia. (3) Severe: meet any of the following: a) respiratory distress, respiratory rate ≥ 30 beats / min; b) in the resting state, means oxygen saturation ≤ 93%; c) arterial blood oxygen partial pressure/ oxygen concentration ≤ 300mmHg (1mmHg = 0.133kPa). Pulmonary imaging showed that the lesions progressed more than 50% within 24-48 hours, and the patients were managed according to moderate case. (4) Critical: one of the following conditions: a) respiratory failure occurs and requires mechanical ventilation; b) Shock occurs; c) ICU admission is required for combined organ failure.

This retrospective study was performed in accordance with the ethical guidelines of the Helsinki Declaration of 1975 and was authorized by the Institutional Review Board of Beijing You’an Hospital, Capital Medical University (Approval No. 2020-022). The informed consent was waived by the IRB because this study was a retrospective assessment.

### Baseline data collection

The laboratory data of 85 patients at admission were collected. Laboratory assessments consisted of complete blood count, blood chemistry, coagulation test, liver and renal function, C-reactive protein, procalcitonin, lactic acid, cardiac markers.

### Statistical analysis

Age and days were represented by median (range), categorical variables by number (%), and laboratory data by mean (interquartile range). Compared the differences between the two groups with t-test, chi-square test or Mann-Whitney U test. Logistic regression was used to select independent risk factors that affect abnormal liver function. Analyses were performed using SPSS 22.0 statistical package (SPSS, Inc., Chicago, IL, USA). P-value < 0.05 was considered statistically significant.

## Results

### Clinical characteristics of patients with COVID-19

The characteristics of patients with COVID-19 in the present study were shown in **Table 1**. The median age of the patients with COVID-19 was 49 years old, the sex ratio was similar. The average ALT value was 28U/L. The average value of total bilirubin and direct bilirubin were in the normal range. The average level of creatinine was normal. C-reactive protein increased significantly, the average value of lymphocyte was close to the normal low value. The median value of creatine kinase was normal. In terms of clinical classification, the proportion of patients with ordinary type was 42.3%, followed by severe and critical patients.

**Table 1.** Characteristics of the patients with COVID-19.

| **Indicators** | **Value** |
| --- | --- |
| Age(years) | 49.0(36.0, 64.0) |
| Sex: Female, n(%) | 38(44.7) |
| ALT(U/L) | 28.0(19.5, 44.5) |
| AST(U/L) | 29.0(20.5, 38.5) |
| Albumin(g/L) | 36.9±5.1 |
| Total bilirubin (μmol/L) | 9.2(6.4, 12.1) |
| Direct bilirubin (μmol/L) | 1.8(1.2, 2.5) |
| Creatinine (μmol/L) | 61.0(53.0, 74.5) |
| Lactic acid(mmol/L) | 1.3±0.6 |
| C-reactive protein(mg/L) | 17.0(2.6, 37.2) |
| White blood cell (10<sup>9</sup>/L) | 4.4(3.5, 5.9) |
| Hemoglobin (g/L) | 133.7±16.1 |
| Platelet (10<sup>9</sup>/L) | 193.0(161.0, 233.0) |
| Neutrophil count(×10<sup>9</sup>/L) | 2.8(1.8, 4.1) |
| Lymphocyte(×10<sup>9</sup>/L) | 1.1(0.7,1.6) |
| Monocyte(×10<sup>9</sup>/L) | 0.3(0.2,0.4) |
| Creatine Kinase(U/L) | 76.0(46.0, 120.0) |
| Creatine Kinase Isoenzyme-MB(U/L) | 0.3(0.2,0.7) |
| Troponin I | 0.01(0.01,0.02) |
| Myoglobin | 40.0(28.5,63.5) |
| INR | 1.1±0.1 |
| History of liver diseases, n(%) | 6(7) |
| **Clinical types , n(%)** |  |
| Mild type | 4(4.7) |
| Ordinary type | 55(64.7) |
| Severe type | 17(20.0) |
| Critical type | 9(10.6) |

### The proportion and incidence rate of abnormal liver function

There were 33 patients with abnormal liver function during the whole process of hospitalization, accounting for 38.8% (33/85), including elevated ALT and AST (**Table 2**). And 24.7% (21/85) of them with ALT elevation on admission. Among the 21 patients presenting with abnormal liver function at admission, only one of them had moderate elevation of ALT (179U/L), others had mild liver injury (ALT<2ULN). 14.1% of patients (12/85) had abnormal liver function during the whole process of hospitalization. All of them are mild and medium elevation of ALT. Six patients with chronic hepatic diseases were enrolled in our study. There was no abnormal liver function in two patients with HBV infection. There were two patients with alcoholic liver disease, one of them had moderate elevation of ALT (120U/L) within one week after admission, the other one had no liver injury. There were two patients with fatty liver, one patient had mild liver injury on admission, the other one had liver injury on 3 days after admission.

**Table 2.** Comparison of baseline data of the COVID-19 patients between normal and abnormal liver function group.

| **Indicators** | **ALT normal group**<br>**(n=52)** | **ALT elevation group**<br>**(n=33)** | **P value** |
| --- | --- | --- | --- |
| Age(years) | 44.5(31.0 , 61.5) | 59.0 (47.0 , 69.5) | 0.001 |
| Sex: Female , n(%) | 29 (57.7) | 18 (54.5) | 0.912 |
| Drinking history, n(%) |  |  | 0.032 |
| Yes | 5 (9.6) | 9 (27.3) |  |
| No | 47 (90.4) | 24 (72.7) |  |
| With other liver diseases<br>, n(%) | 3(5.8) | 3(9.1) | 0.882 |
| Ibuprofen, n(%) |  |  | 0.714 |
| Yes | 5 (9.6) | 4 (27.3) |  |
| No | 47 (90.4) | 29 (72.7) |  |
| Albumin(g/L) | 37.8±4.8 | 33.4±4.3 | 0.000 |
| Total bilirubin (μmol/L) | 8.7±3.7 | 11.8±5.3 | 0.003 |
| Direct bilirubin (μmol/L) | 1.6±0.7 | 3.7±1.7 | 0.001 |
| Creatinine (μmol/L) | 60.0 ( 51.5 , 74.0 ) | 65.0 ( 56.0 , 77.5 ) | 0.267 |
| Lactic acid(mmol/L) | 1.2±0.5 | 1.5±0.6 | 0.037 |
| C-reactive protein(mg/L) | 7.4 ( 1.7 , 21.3 ) | 39.7 ( 16.2 , 70.6 ) | 0.000 |
| White blood cell<br>(*10<sup>9</sup>/L) | 4.3 ( 3.5 , 5.8 ) | 5.1 ( 3.5 , 6.6 ) | 0.332 |
| Hemoglobin (g/L) | 132.6±17.0 | 135.4±14.6 | 0.439 |
| Platelet (*10<sup>9</sup>/L) | 207.0 ( 161.0 , 250.3 ) | 180.0 ( 160.0 , 211.0 ) | 0.189 |
| Neutrophil count(*10<sup>9</sup>/L) | 2.3 ( 1.5 , 3.5 ) | 3.2 ( 2.3 , 5.0 ) | 0.006 |
| Lymphocyte<br>count(*10<sup>9</sup>/L) | 1.4 ( 0.9 , 1.8 ) | 0.8 ( 0.6 , 1.2 ) | 0.000 |
| Monocyte count(*10<sup>9</sup>/L) | 0.3 ( 0.2 , 0.4 ) | 0.3 ( 0.2 , 0.3 ) | 0.076 |
| Creatine Kinase(U/L) | 69.5 ( 45.3 , 103.3 ) | 89.0 ( 46.5 , 139.0 ) | 0.246 |
| Creatine Kinase<br>Isoenzyme-MB(U/L) | 0.31 ( 0.15 , 0.71 ) | 0.38 ( 0.21 , 0.80 ) | 0.525 |
| TroponinI | 0.01 ( 0.01 , 0.02 ) | 0.01 ( 0.01 , 0.02 ) | 0.412 |
| Myoglobin | 33.5 ( 26.5 , 54.8 ) | 53.0 ( 34.0 , 112.5 ) | 0.001 |
| INR | 1.1±0.1 | 1.2±0.1 | 0.117 |
| Clinical classification, n (%) |  |  | 0.017 |
| Mild and Ordinary type | 42(80.7) | 17(51.5) |  |
| Severe type | 7(13.5) | 10(30.3) |  |
| Critical type | 3(5.8) | 6(18.2) |  |

### Comparison of baseline data of the COVID-19 patients between normal and abnormal liver function group

The results were shown in **Table 2**. The median age of the two groups was statistically different, the ALT elevation group was 59 years old and the ALT normal group was 44.5 years old. Compared with patients with normal liver function, patients who had elevated ALT were more likely to have drinking history. Albumin in ALT elevation group were significantly lower than those in the ALT normal group (33.4±4.3 vs 37.8±4.8, *P* = 0.000). The bilirubin level was normal between the two groups. The level of lactic acid was significantly higher in the ALT elevation group (1.5±0.6 vs 1.2±0.5, *P* = 0.037). The number of lymphocytes in ALT elevation group was significantly lower than that in ALT normal group (0.8 vs 1.4, *P* = 0.000). However, the level of CRP was significantly higher in the ALT normal group (39.7 vs 37.4, *P* = 0.000). At the same time, the number of neutrophils and myoglobin also increased in ALT elevation group than in ALT normal group. The proportion of severe and critical patients in the ALT elevation group was significantly higher, reaching 61.5%.

### The factor related to ALT elevation in Multivariate analysis

Multivariate logistic regression was performed to analyze the factors including age, drinking history, baseline albumin, lactic acid, CRP, neutrophils, lymphocytes, myoglobin and clinical classification. It was found that the CRP ≥20 mg/L and lymphocyte count< 1.1×10^9/L were independently related to ALT elevation. (**Table 3**)

**Table 3:** Multivariate analysis of COVID-19 patients with abnormal liver function.

| Indicators | OR (95%CI) | P value |
| --- | --- | --- |
| C-reactive protein level $\geq 20$ mg/L | 3.206(1.262~8.144) | 0.014 |
| Lymphocyte count $< 1.1 \times 10^9$/L | 0.263(0.104~0.667) | 0.005 |

*OR: Odds ratio, CI: Confidence interval*

## Discussion

The present study mainly focused on the risk factors involved with hepatic injury in the patients with COVID-19. After excluding the patients receiving drugs that may cause liver damage such as lopinavir/ritonavir, according to the findings, for the first time, we found that the level of lymphopenia and CRP were independently associated with hepatic injury, and our study suggested inflammatory cytokine storm might be the major mechanism.

As the number of patients with COVID-19 continues to rise, liver injury is frequently reported as extra-pulmonary clinical feature. Some recent studies revealed that nearly a half of patients experienced different degrees of liver injury^7.8^. However, as far as we know, there is no study on the factors of liver injury. Such as SARS, SARS-CoV-2 uses the same cell entry receptor angiotensin converting enzyme-II^4^, which is expressed in the liver tissue as well. A lately research showed that bile duct cells had higher specific expression of angiotensin converting enzyme-II than hepatocytes^12^, which indicated the possibility of SARS-CoV-2 infected bile duct cells directly and caused hepatic injury. It’s suggested that bile duct epithelial cells may play a key role in immunoreaction. However, clinical data from two researches showed that the bile duct injury related indexes alkaline phosphatase, gamma-glutamyl transpeptidase and total bilirubin did not increase significantly^10,14^. In our study, the hepatic injury of patients was mainly reflected through mild or medium elevation of ALT, and the level of serum TBIL was almost normal. Moreover, the pathological results of liver biopsy showed moderate microvascular steatosis and active inflammation in the hepatic lobule portal area^13^. Due to the use of lopinavir/ritonavir during treatment, it is difficult to distinguish the reason of liver injury according to the pathological results. Although the cell entry receptor angiotensin converting enzyme-II is highly expressed in bile duct cells, recent works and our findings suggested that SARS-CoV-2 infection did not cause an obvious bile duct injury. Thus, SARS-COV-2 infection may not be the major reason related to liver injury.

In our study, there were a total of 33 patients with abnormal liver function during the whole process of hospitalization, accounting for 38.8%. And 24.7% (21/85) of them showed ALT elevation on admission. Among the 21 patients presenting with hepatic injury, only one of them had moderate elevation of ALT (179U/L), others had mild liver injury (ALT<2ULN). The results suggested that COVID-19-related hepatic injury was not serious at the beginning of the disease, which was consistent with previous researches. Compared with other study^9^, the higher proportion of patients with liver injury at admission might be related to the number of samples and the different periods from onset to hospitalization in our study.

Among the patients presenting with abnormal liver function in the present study, moderate and severe types of patients were more likely to have liver injury, accounting for 58.8% and 66.7%. This indication was consistent with Academician Nanshan Zhong ‘s recent clinical research on acute respiratory syndrome caused by COVID-19, which involved the largest scale of data so far^15^. It suggested that hepatic injury likely occurred in patients under critical care. Moreover, in our study, a positive correlative relationship between the lymphopenia and liver injury. Based on previous studies^16,17^, lymphopenia is typical laboratory indicator during highly pathogenic coronavirus infections, such as the SARS-CoV and MERS-CoV infection, and is believed to be associated with disease severity. Recent studies revealed that 63%-70.3% of SARS-CoV-2 infected patients with severe diseases have lymphopenia and the low level of lymphocyte counts in patients is associated with mortal outcome^8,18^. Therefore, our results suggested that the occurrence of hepatic injury was related to the deterioration of the disease with a dynamic process. In addition, six cases with chronic hepatic diseases were enrolled in our study, however there was no significant difference between the abnormal liver function group and normal liver function group. This might be related to the fact that most of the inpatients were ordinary type^9,10^.

Previous studies have shown that inflammatory cytokine storm is associated with severe lung injury and adverse outcomes of SARS-CoV or MERS-CoV infection ^17,19,20^. Inflammatory cytokine storm is an overactive inflammatory response of the human body caused by virus infection, which leads to a persistent activation and reproduction of lymphocytes and macrophages that will secrete huge amount of inflammatory cytokine^21^. Inflammatory cytokine storm not only leads to pulmonary injury, but also the injury of non-pulmonary organs including the liver, kidneys and cardiac muscle. And some studies have found the liver damage is more common in patients with severe pneumonia, which is suspected to be associated with inflammatory factor storm^8,9^. In our study, patients with liver damage had higher elevated CRP level. As a typical inflammatory index, CRP is an acute phase protein that rapidly increase in the presence of infection or tissue damage. Additionally, lymphocyte cells are important for inhibiting overactive innate immune responses during viral infection^22,23^. Thus, loss of lymphocyte cells during SARS-CoV-2 infection may result in aggravated inflammatory responses. The significant decreases in the counts of lymphocyte cells, especially CD8^+^T cells, as well as increases in IL-6, IL-10, IL-2 and IFN-γ levels in the peripheral blood of patients with COVID-19^24^. While T cell counts drop to the lowest levels, serum level of inflammatory cytokines reach their peaks in a few days after the onset of COVID-19. It suggests that inflammatory cytokines are involved in the process at the early stage, which can explain the reason for the patients with mild liver injury.

Compared with the inflammatory damage of lung, the inflammatory damage of liver was relatively mild by autopsy analysis^13^. This result may explain that no one develop to liver failure or severe hepatitis in our study, even in severe cases. 14.1% of patients (12/85) had liver injury during the whole process of hospitalization. All of them are mild and medium elevation of ALT. After liver protection treatment, the liver function of most patients was improved within one or two weeks.

Our study also observed the patients had a decline of serum albumin, though there was no obvious difference between the two groups. It was more likely due to the patients’ hyper metabolic state presenting with fever, malnutrition and low caloric intake, suggesting the importance of nutritional support therapy. Moreover, hyperlactatemia reflects hypoxia injury and may be related to liver injury, our results showed that hyperlactatemia was related to liver injury, but it turned to negative after multivariate analysis. The change of result may be related to our sample size, which needs further study and confirmation.

There were some limitations in our study. The data were collected from a single center at a certain timepoint. So, the sample size was relatively limited. And large-sample cohort studies are still needed to verify this in the future.

In conclusion, hepatic injury is likely a complication of COVID-19 infection. Inflammatory cytokine storm may be the major mechanism. Early detection and timely treatment are helpful for the recovery of liver function.

## Data Availability

All data referred to in the manuscript are available.

## Acknowledgments

This study was funded by National Science and Technology Key Project on “Major Infectious Diseases such as HIV/AIDS, Viral Hepatitis Preventon and Treatment” (NO. 2017ZX10203201-005), Beijing Municipal Administration of Hospitals Clinical Medicine Development of Special Funding Support (NO. ZYLX201806), National Key R&D Program of China (No.2017YFA0103000), Medical Science Research Project Support by Bethune Charitable Foundation

